# Spatial, Temporal, and Molecular Heterogeneity of ADC targets in High-Grade Serous Ovarian Carcinoma

**DOI:** 10.64898/2025.12.05.25341695

**Authors:** Xiaoxuan Li, Tobias Janik, Markus Möbs, Stefan Florian, Wolfgang D. Schmitt, Amra Dzakulic, Jalid Sehouli, David Horst, Frank Dubois, Ioana Elena Braicu, Mihnea P. Dragomir

## Abstract

**Background:** Antibody-drug conjugates (ADCs) represent a promising therapeutic approach for high-grade serous ovarian carcinoma (HGSOC). Patient selection for ADC therapy depends on tumor target expression, making it essential to characterize molecular, spatial, and temporal heterogeneity.

**Methods:** We analyzed two HGSOC tissue microarray cohorts: 100 genomically profiled primaries (1,565 cores) and 64 matched cases with paired primary (P), primary disseminated (PD), and recurrent (R) samples (2,395 cores). Associations between ADC target expression and molecular characteristics, sampling site, and survival were investigated.

**Results:** ADC targets showed no significant associations with homologous recombination deficiency (HRD) or *TP53* mutation status; TROP2 was modestly lower in *BRCA1/2*-mutated tumors. Folate receptor-alpha (FolR1) showed notable spatial heterogeneity: 25% switched therapeutic-indication groups between center and margin at P; 21.7% were reclassified between P and PD. Temporally, all markers showed ≥20% switching, reaching 38.4% for FolR1 between P and R. High FolR1 expression in P correlated with poorer survival, a pattern not observed in PD or R samples.

**Conclusions:** ADC targets in HGSOC display limited molecular but significant spatial and temporal heterogeneity, with expression classifications varying by site and time. FolR1 expression in primary tumors associates with aggressive disease.

## 1. Introduction

Ovarian cancer is the leading cause of death from gynecological malignancies worldwide. High-grade serous ovarian carcinoma (HGSOC) is the most common and aggressive type with a poor prognosis^1^. Its high recurrence rate and treatment resistance are major problems in its clinical management^2^. One of the most characteristic molecular alterations in HGSOC is *TP53* mutation, present in nearly all cases^3–6^. In addition, HGSOC shows marked molecular heterogeneity. A major component of this heterogeneity is impaired DNA repair capacity, most notably homologous recombination deficiency (HRD), the loss of the homologous recombination repair (HRR) pathway for DNA double-strand break repair^7,8^. About half of HGSOCs are HRD-positive, most commonly due to *BRCA1/2* mutations^9,10^. This subgroup is particularly sensitive to platinum-based chemotherapy and PARP inhibitors, and HRD status is associated with improved overall survival (OS), even without PARP inhibition^11–17^. Beyond genomic alterations, HGSOC also exhibits spatial and temporal heterogeneity at the histological level, with site-specific variations observed between the primary ovarian tumor and peritoneal metastases^18,19^.

Antibody–drug conjugates (ADCs) deliver cytotoxic agents to tumor cells via monoclonal antibodies. Folate receptor alpha (FRα/FolR1) is highly expressed in HGSOC but largely absent in normal tissues, making it an attractive therapeutic target ^20–22^. Mirvetuximab soravtansine (MIRV), an ADC targeting FolR1, is EMA- and FDA-approved for platinum-resistant ovarian cancer, underscoring the clinical relevance of this strategy^20,23^. Beyond FolR1, other antigens have emerged as promising targets for ADCs. Trophoblast cell-surface antigen 2 (TROP2) is frequently overexpressed in HGSOC and linked to poor prognosis, with the TROP2-targeted ADC datopotamab deruxtecan (Dato-DXd) showing potent preclinical and clinical efficacy ^24–27^. HER2, although less common in HGSOC, correlates with aggressive tumor behavior; HER2-directed ADCs such as trastuzumab emtansine (T-DM1) and trastuzumab deruxtecan (T-DXd), have shown antitumor activity in preclinical ovarian cancer models, with the last one receiving tumor agnostic FDA approval in Her2 2+/3+ expressing tumors^28,29^. Nectin-4, a tumor-associated antigen highly expressed in cancer cells but largely absent in normal adult tissues, has been linked to adverse prognosis in ovarian cancers^30–36^. It represents a promising ADC target, exemplified by the approval of enfortumab vedotin for advanced urothelial carcinoma^37,38^; however, its role in ovarian cancer remains unclear. The selective expression of FolR1, TROP2, HER2, and Nectin-4 highlights their therapeutic potential as ADC targets in HGSOC and provides a strong rationale for systematic evaluation. Importantly, patient eligibility is typically defined by immunohistochemistry (IHC) thresholds, such as FolR1-high status in MIRV trials and HER2 scoring in ongoing studies.

Although ADC targets show promise in ovarian cancer, most studies have focused on single antigens without systematic comparison under uniform conditions. Key issues remain unresolved, including their overall expression frequencies, inter-marker correlations, links to the molecular heterogeneity of HGSOC, and potential spatial or temporal variation across anatomical sites, tumor regions, or disease progression. Another gap is whether IHC-defined thresholds of ADC targets are associated with prognosis and reliably guide patient stratification. These limitations underscore the need for an integrated assessment of ADC target heterogeneity in HGSOC.

We hypothesize that ADC targets in HGSOC display molecular heterogeneity, interacting with genomic features such as HRD status, *TP53*, and *BRCA1/2* mutations to influence tumor biology and outcomes. We further hypothesize that ADC target expression also varies spatially and temporally, across primary ovarian and primary dissemination sites, tumor regions, and between primary and recurrent diseases. Such heterogeneity may affect biomarker evaluation and patient stratification for ADC therapy.

## 2. Materials and Methods

### 2.1 Study Cohorts

This study included two non-overlapping cohorts comprising 164 HGSOC cases who underwent surgery at Charité University Hospital Berlin or affiliated hospitals between 2014 and 2024 (Fig. 1). All tumors were histopathologically reviewed, classified according to the World Health Organization (WHO) Classification of Female Genital Tumors (5th Edition), and staged by the American Joint Committee on Cancer (AJCC) Cancer Staging Manual (8th Edition).

**Figure 1.**
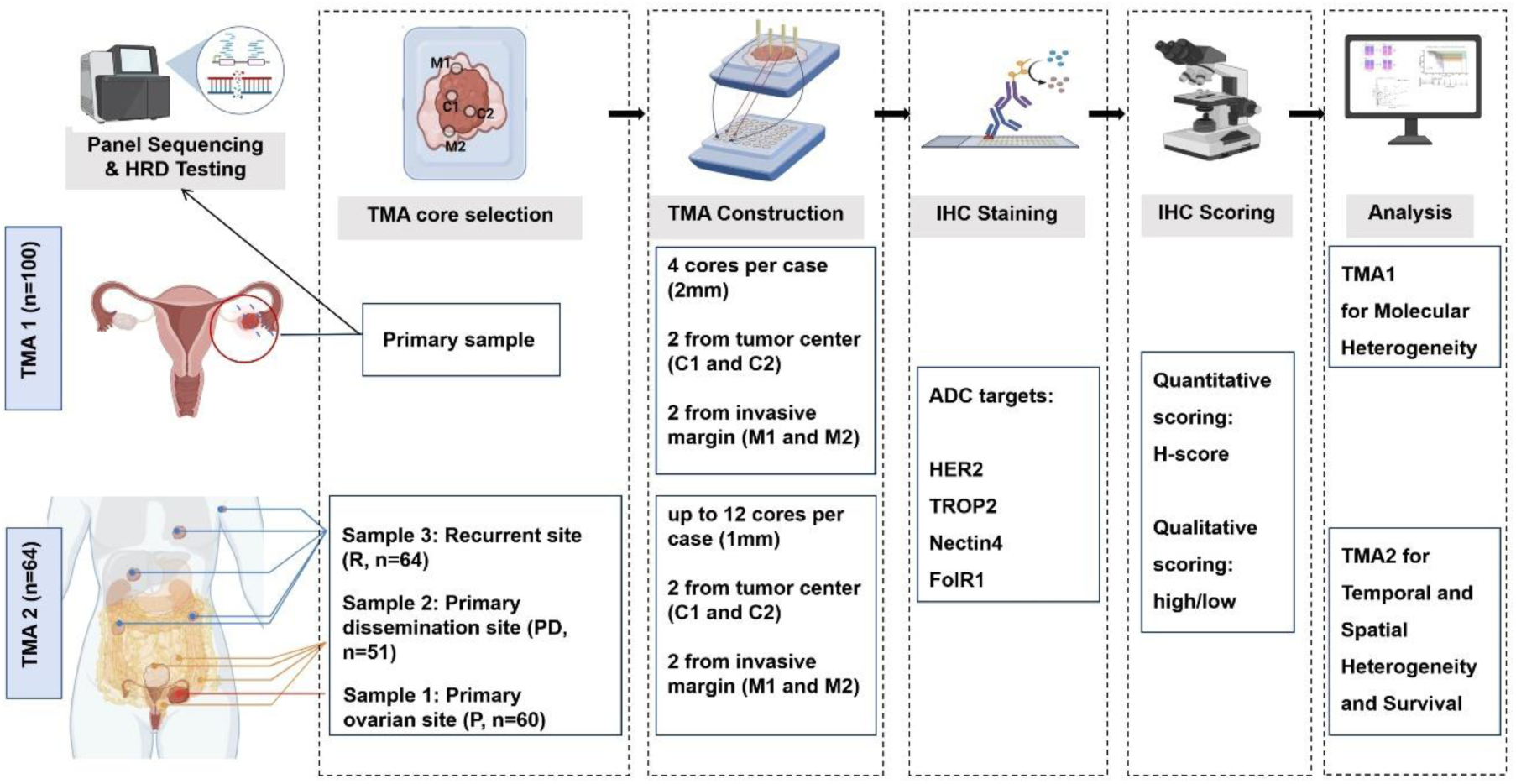
Overview of the study design, tissue microarray (TMA) construction, and the analytical workflow. TMA1 included 100 primary HGSOC cases, with four 2-mm cores per case sampled from the tumor center (C1, C2) and invasive margin (M1, M2). All cases underwent panel sequencing and HRD testing. TMA2 comprised 64 recurrent HGSOC cases, each with up to three tumor sites: primary ovarian tumor (Sample 1, P), primary dissemination (Sample 2, PD), and recurrent tumor (Sample 3, R). Each site contributed four 1-mm cores, resulting in up to 12 cores per case. All samples were stained for four ADC targets (HER2, TROP2, Nectin-4, and FolR1) and evaluated using both H-score and high/low classification. The graphic elements were created on Biorender.com.

Cohort 1 (TMA1) consisted of 100 primary HGSOC cases reviewed and tested for homologous recombination deficiency (HRD) at the Institute of Pathology, Charité, between 2021 and 2024. HRD testing was performed using the NOGGO GIS v1 assay (details in Section 2.4). Cohort 2 (TMA2) comprised 64 HGSOC cases diagnosed between 2014 and 2021, all of which recurred between 2015 and 2023. Each patient underwent surgery and tumor sampling at both primary diagnosis and recurrence. For both cohorts, clinical and pathological data were collected, including age at diagnosis, T stage, FIGO stage, follow-up information, and detailed treatment history (Supplementary Table S1).

The conduct of this study was approved by the local ethics committee (EA1/110/22).

### 2.2 Tissue Microarray (TMA) Construction

For both cohorts, formalin-fixed, paraffin-embedded (FFPE) tumor blocks were used to construct two independent TMAs. Lymphoid tissue was included on each array for orientation. Cohort 1 (TMA1) comprised 100 primary HGSOC cases, each represented by four 2.0-mm cores: two from the tumor center (C1, C2) and two from the invasive margin (M1, M2). In total, 1565 valid cores were available for analysis. Cohort 2 (TMA2) included 64 HGSOC patients with matched recurrent tumors. Based on sample availability, three tumor sites were represented: primary ovarian tumor (hereafter P, n=60), primary dissemination (most often greater omentum, hereafter PD, n=51), and recurrence (Sample 3, hereafter Recurrent [R], n=64). From each site, four 1.0-mm cores were sampled (two center [C1, C2] and two margin [M1, M2]). Of the 64 cases, 47 had cores from all three sites. Tissue loss occurred in one case during Nectin-4 and FolR1 IHC; four cases lacked P but had PD, and 13 lacked PD but retained P. In total, 2395 valid cores were available from TMA2. Altogether, 3960 evaluable tissue cores were included in the study. The analytical workflow, including TMA construction, IHC staining and scoring, and statistical analysis, is illustrated in Figure 1.

### 2.3 Immunohistochemistry (IHC) and Scoring

For IHC, TMA blocks were cut into 2-µm sections and incubated in CC1 buffer (Ventana) according to protocol (CC1 mild for HER2/Nectin-4, CC1 standard for FolR1, 16-min CC1 for TROP2). Primary antibodies were anti-HER2 (clone 4B5, Ventana, RTU), anti-Nectin-4 (clone EPR15613-68, Abcam, 1:100), anti-TROP2 (clone 01, Enzo, 1:500), and anti-FolR1 (clone FOLR1-2.1, Roche, RTU). Incubation was 32 min at room temperature, with visualization by the avidin-biotin complex method and 3,3′-diaminobenzidine. Staining was performed on the BenchMark Ultra platform (Ventana). Nuclei were counterstained with hematoxylin/bluing reagent (Ventana) for 12 min. Slides were evaluated using Olympus BX50/BX46 microscopes, and whole-slide images were acquired with a high-resolution digital scanner for pathological review.

IHC staining was primarily assessed by a certified pathologist (X.L.); cases with uncertain scores were independently reviewed by two other pathologists (F.D. and M.P.D.), and results were reached by consensus. For ADC-target markers, staining intensity was graded as 0 (negative), 1+ (weak), 2+ (moderate), or 3+ (strong). The percentage of tumor cells at each level was recorded, and an H-score was calculated: H-score = (% of 1+ cells) × 1 + (% of 2+ cells) × 2 + (% of 3+ cells) × 3, ranging from 0–300.

For ADC marker classification, binary thresholds were applied based on previous studies or clinical trial data. HER2 expression was evaluated according to the American Society of Clinical Oncology and the College of American Pathologists (ASCO/CAP) guidelines for gastroesophageal adenocarcinoma^39^. A score of 3+ was defined as strong membranous staining, either complete or basolateral, in ≥10% of tumor cells; 2+ indicated moderate to weak staining in ≥10% of cells; 1+ weak or incomplete staining in ≥10%; and 0 no staining or <10% of cells. For statistical purposes, HER2 scores of 2+ and 3+ were defined as high expression, while 0 and 1+ were defined as low expression^40^. For TROP2, we used an H-score cutoff of 200, as reported in recent OC/EC studies^26^. H-score ≥200 defined high, <200 low. FolR1 high expression was defined as ≥75% of tumor cells with moderate/strong staining^41^. Nectin-4 was classified as 0 (H-score 0–14), 1+ (15–99), 2+ (100–199), and 3+ (200–300)^42^; H-scores ≥100 were considered high, <100 low. Representative IHC staining patterns and scoring criteria for each ADC target are illustrated in Figure 2.

**Figure 2.**
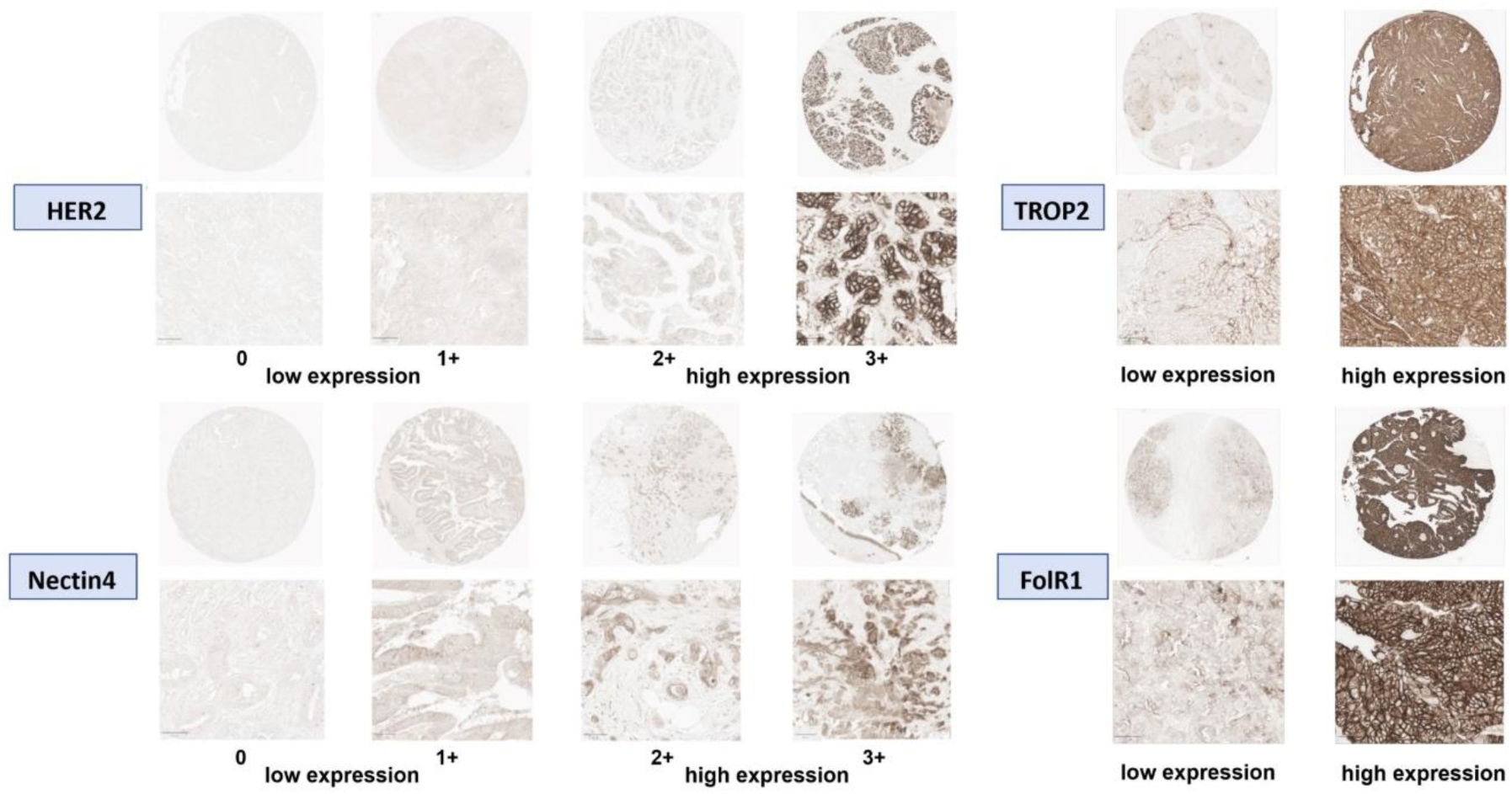
Representative IHC expression patterns and scoring criteria for ADC targets. HER2 and Nectin-4 were scored using a 4-tier system (0, 1+, 2+, 3+); for binary classification, scores of 0 and 1+ were considered low expression, and 2+ and 3+ as high expression, as defined in the Methods section. TROP2 and FolR1 were assessed using a binary classification system (low vs. high).

### 2.4 Molecular Profiling (Cohort 1)

Homologous recombination deficiency (HRD) was assessed with the NOGGO GIS v1 assay^43^, a hybrid-capture NGS panel covering *BRCA1/2*, 55 HRR genes, additional driver mutations (*TP53*, *KRAS*, *NRAS*, *PIK3CA*, *BRAF*), and copy number alterations. Genomic instability (GI) was measured by three parameters: percentage of loss of heterozygosity (PLOH), copy number alterations (PCNA), and telomeric copy number alterations (PTCNA), which were combined to generate a genomic instability score (GIS). HRD positivity was defined as either a GIS ≥83 or the presence of pathogenic *BRCA1/2* mutations. *TP53* mutations were classified as missense, in-frame indels, splice-site alterations, or truncating mutations (including nonsense and frameshift). For *TP53*, missense/in-frame variants were analyzed together because they usually produce full-length p53 with dominant-negative or gain-of-function properties. In contrast, splice-site and truncating variants disrupt canonical splicing or introduce premature stop codons, which typically trigger nonsense-mediated decay or produce truncated proteins lacking critical domains, resulting in loss of function^44^. We therefore combined variants by their expected protein consequence to increase statistical power while reflecting mechanistic similarity.

### 2.5 Statistical Analysis

Analyses were performed in R (v4.4.2). All tests were two-sided, with p<0.05 considered significant.

#### 2.5.1 Molecular Heterogeneity Analysis in Cohort 1

To evaluate associations between ADC target expression and HRD, *TP53*, and *BRCA1/2* status, we compared quantitative H-scores (non-normal by Shapiro–Wilk) using Wilcoxon rank-sum tests and high/low categories using χ² or Fisher’s exact tests. For *TP53*, missense and in-frame variants were grouped, and splice-site variants with truncating variants, reflecting functional similarities. For *BRCA1/2* as mutated versus wild type.

#### 2.5.2 Spatial Heterogeneity Analysis in Cohort 2

We compared margin (M) vs center (C) at P, PD, and R. With two cores per region (M1/M2, C1/C2), we used the mean H-score and assigned the higher category if categorical calls differed; single cores were used as is. Pairs require 1 M and ≥1 C core. Wilcoxon signed-rank tested H-scores; McNemar tested high/low. Results were checked for replication in Cohort 1. We also assessed P vs PD pairs using the same methods.

#### 2.5.3 Temporal Heterogeneity Analysis in Cohort 2

We assessed temporal heterogeneity of ADC targets in Cohort 2 by comparing expression between paired primary and recurrent tumors (P–R and PD–R). H-scores were analyzed with Wilcoxon signed-rank tests; categorical high/low with McNemar tests. Sankey plots visualized category changes. The switching rate between groups was defined as the proportion of cases that changed category (low-high) from P/PD to R.

#### 2.5.4 Survival Analysis in Cohort 2

Survival analysis was conducted in Cohort 2, which possessed complete longitudinal follow-up data of sufficient duration. Overall survival (OS) was defined as the interval from the date of primary diagnosis to death or last follow-up. Progression-free survival (PFS) was defined as the interval from the date of primary diagnosis to radiologically or clinically confirmed progression or death, whichever occurred first. Post-recurrence survival (PRS) was defined as the interval from the date of tissue sampling at recurrence to death or last follow-up. Classical PRS from first recurrence could not be calculated because biopsy timing did not uniformly coincide with the first recurrence event. We assessed the prognostic value of ADC targets by Kaplan–Meier curves: OS and PFS in P/PD tumors and PRS in R tumors, comparing high vs low expression with the log-rank test. Associations with survival were further tested using univariable Cox models; significant markers were entered into multivariable models with clinical covariates. Age, T-stage, and platinum sensitivity were selected as covariates, as they are established prognostic factors in HGSOC. Platinum sensitivity was defined by the platinum-free interval (PFI): <6 months resistant, 6–12 months partially sensitive, and >12 months sensitive^45,46^. Restricting the model to these variables reduces the risk of overfitting and ensures that the independent prognostic value of ADC marker expression is evaluated within the context of the most relevant clinical parameters.

## 3. Results

### 3.1 Most HGSOCs show expression of multiple ADC targets without any correlation to known molecular subtypes

To test our hypothesis that ADC target expression reflects molecular heterogeneity in HGSOC and interacts with genomic features such as *HRD*, *TP53*, and *BRCA1/2*, we analyzed 100 genomically well-characterized primary tumors.

Across this cohort (n = 100; 1,565 evaluable TMA1 cores), ADC marker expression varied widely. TROP2 (75%) and FolR1 (56%) were most frequently overexpressed, whereas HER2 (34%) and Nectin-4 (36%) showed lower prevalence. Among HER2-high cases, most (31/34) exhibited 2+ intensity, while only a minority (3/34) reached 3+ intensity (Fig. 3A). We assessed whether ADC target expression differed by HRD status. Quantitative H-scores did not significantly differ between HRD-positive and HRD-negative tumors (all p > 0.05, Wilcoxon test; Supplementary Fig. S1A). Likewise, the prevalence of high versus low expression showed no significant association with HRD (all p > 0.05, Chi-square test; Fig. 3B).

**Figure 3.**
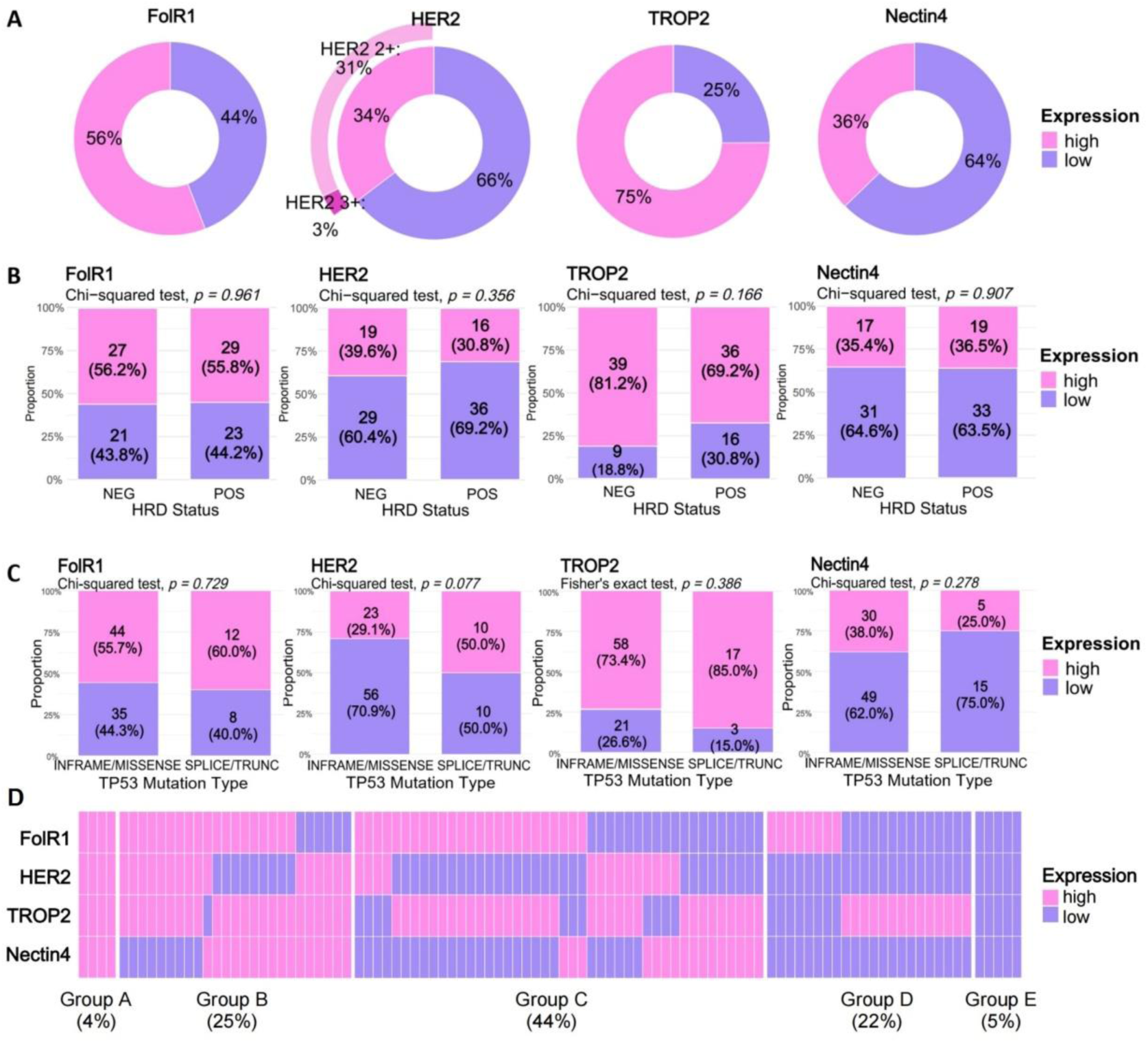
Limited molecular ADC heterogeneity in Cohort 1. (**A**) Distribution of high and low expression levels for FolR1, HER2, TROP2, and Nectin4 across Cohort 1. (**B**) Proportion of ADC target expression groups stratified by HRD status. **(C)** Proportion of ADC target expression groups stratified by *TP53* mutation type (in-frame/missense vs splice-site/truncating). **(D)** Co-expression groups based on the number of high-expressed ADC targets (Group A: all targets high expression; Group B: 3 targets high expression; Group C: 2 targets high expression; Group D: 1 target high expression; Group E: all targets low expression). **Abbreviations**: HRD, homologous recombination deficiency; NEG, negative; POS, positive.

Stratified by *BRCA1/2* status, TROP2 H-scores were modestly lower in mutated than wild-type tumors (Wilcoxon p=0.0289), whereas FolR1, HER2, and Nectin-4 showed no clear differences (Supplementary Fig. S1B). By *TP53* mutation type, none of the four markers differed in H-score (Supplementary Fig. S1C). By *BRCA1/2* status, high-expression rates were broadly similar—HER2 34.7% vs 32.0%, TROP2 77.3% vs 68.0%, Nectin-4 34.7% vs 40.0%, FolR1 52.0% vs 68.0%—with no significant differences (Supplementary Fig. S2).A borderline trend was noted for HER2 (χ² = 3.80, p = 0.077), with higher expression more frequent in splice-site/truncating *TP53* mutations compared to in-frame/missense mutations (Fig. 3C).

Next, we investigated whether the four ADC targets were co-expressed within individual tumors. Spearman correlation analysis of H-scores showed only a weak positive correlation between HER2 and Nectin-4 (ρ = 0.20, p = 0.0435) (Supplementary Fig. S3). We next summarized tumors by the number of high-expression targets. In Cohort 1, 5% of cases were antigen-negative (0/4 high); 22% showed a single target only, whereas 73% showed ≥2 high-expressed targets (Fig. 3D).

Taken together, these results indicate that in primary HGSOC, ADC target H-scores show no significant differences by HRD or *TP53* status, with a modest *BRCA1/2*–TROP2 association (lower TROP2 in *BRCA1/2*-mutated vs wild type). Co-expression was limited, with only a weak HER2–Nectin-4 correlation observed. Nevertheless, most cases (73%) still exhibited more than one high-expression target, indicating that patients are not limited to a single ADC option, and that sequential or alternative targeting strategies remain feasible.

### 3.2 ADC Targets show site-specific heterogeneity leading to discrepant eligibility scores

In Cohort 2 (n = 64; 2,395 evaluable TMA2 cores), a similar pattern of ADC expression was observed in the primary ovarian site (P): TROP2 (85%) and FolR1 (57%) remained the most prevalent high-expression targets, whereas Nectin-4 (37%) and HER2 (20%) were less common. To test our hypothesis that ADC targets display spatial heterogeneity within individual tumors and between primary sites, we compared marker expression between the tumor center (C) and invasive margin (M) at the primary (P), primary dissemination (PD), and recurrent (R) sites, and between P and PD.

First, we compared the margin versus center in primary ovarian tumors (P). At P, paired H-scores showed no M–C difference for any marker (all p>0.3). At PD, only Nectin-4 showed an M–C difference (p=0.0038), whereas FolR1, HER2, and TROP2 did not. At R, no significant M–C differences were detected (all p>0.2) (Supplementary Fig. S4). We then validated M–C comparisons in primary-only Cohort 1: Nectin-4 was not significant (p=0.150), whereas HER2 showed a significant shift between margin and center (p=0.0049; r=0.284; Supplementary Fig. S5). FolR1 showed the highest proportion of switching between therapeutic-indication groups, more specifically switching between high expression group and low expression group or *vice versa* (25.0%; McNemar p=0.4240), followed by TROP2 17.9% (p=0.1084), Nectin-4 19.6% (p=1.0000), and HER2 7.2% (p=1.0000) (Fig. 4A). In PD and R, switching between M and C was likewise appreciable: FolR1 20.0% (PD, p=1.0000) and 9.3% (R, p=1.0000); TROP2 11.1% (PD, p=0.3750) and 24.5% (R, p=0.2668); Nectin-4 15.5% (PD, p=0.1250) and 20.4% (R, p=0.2268); HER2 11.1% (PD, p=1.0000) and 11.2% (R, p=1.0000) (Fig. 4A). Overall, ADC target expression was consistent between M–C, and if significant, the effects were inconsistent across cohorts, arguing against a reproducible center–margin difference.

**Figure 4.**
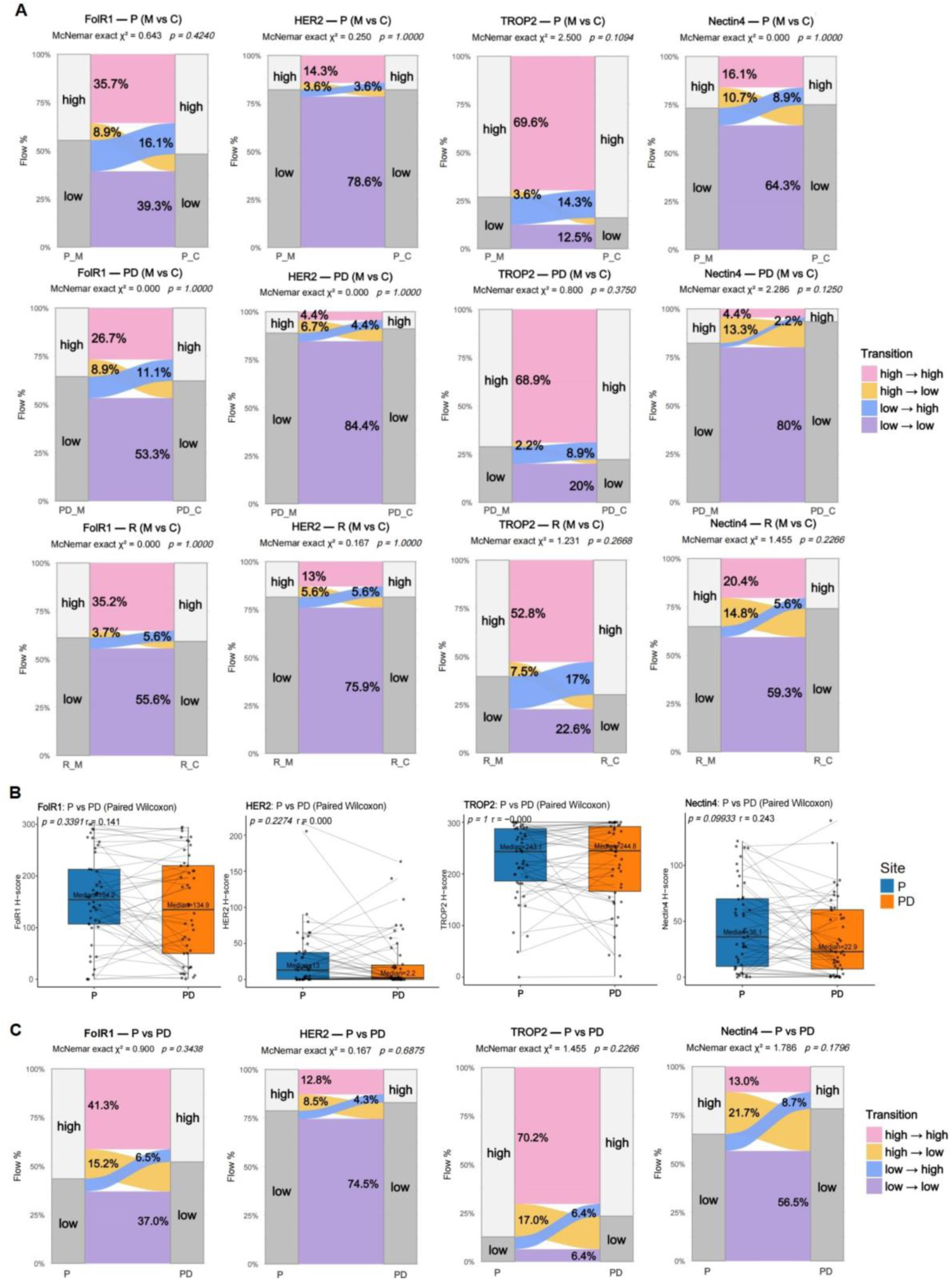
Spatial heterogeneity of ADC target expression in Cohort 2. **(A)** Sankey plots showing categorical transitions between tumor margin (M) and center (C) at P, PD, and R across four ADC markers: FolR1, HER2, TROP2, and Nectin-4 (left to right). **(B)** Paired H-score comparisons between P and PD for the four markers. Statistical significance was assessed using the paired Wilcoxon test. **(C)** Sankey plots showing categorical group transitions in paired PD and R samples across four ADC markers: FolR1, HER2, TROP2, and Nectin4 (left to right). **Abbreviations**: P, primary ovarian site; PD, primary dissemination; R, recurrent tumor; C, tumor center; M, tumor margin.

Second, we analyzed spatial heterogeneity by comparing the ADC target expression between matched P samples and PD samples. H-scores showed no significant differences (FolR1 p=0.3391; HER2 p=0.2274; TROP2 p=1.000; Nectin-4 p=0.0993; Fig. 4B). Nevertheless, switching between therapeutic-indication groups occurred in a notable fraction of cases: FolR1 showed substantial reclassification (21.7%; p=0.3438); switching in the other markers was 12.8% for HER2 (p=0.6875), 23.4% for TROP2 (p=0.2266), and 30.4% for Nectin-4 (p=0.1796) (Fig. 4C).

ADC eligibility hinges on binary IHC classifications yet switching between therapeutic-indication groups varies by sampling site and region. For FolR1, a 25% margin–center discordance at P and 21.7% P–PD reclassification indicates that single-site/region assessments can alter eligibility. We therefore recommend standardized multi-site/region sampling, explicit site/region documentation, and targeted IHC re-testing at decision points.

### 3.3 ADC targets expression frequently switches between primary and recurrent tumors

To evaluate our hypothesis that ADC targets undergo temporal changes during disease progression, first, we compared primary ovarian tumors (P) with matched recurrent tumors (R). H-scores increased for Nectin-4 in R (p=0.0134; r=0.319), while HER2, TROP2, and FolR1 showed no significant change (p=0.1375, 0.2279, 0.2575, respectively) (Fig. 5A). For binary IHC classifications, switching between therapeutic-indication groups was substantial: FolR1 38.4% (McNemar p=0.6776), Nectin-4 40.0% (p=0.5413), TROP2 26.7% (p=0.4545), and HER2 20.0% (p=1.0000).

**Figure 5.**
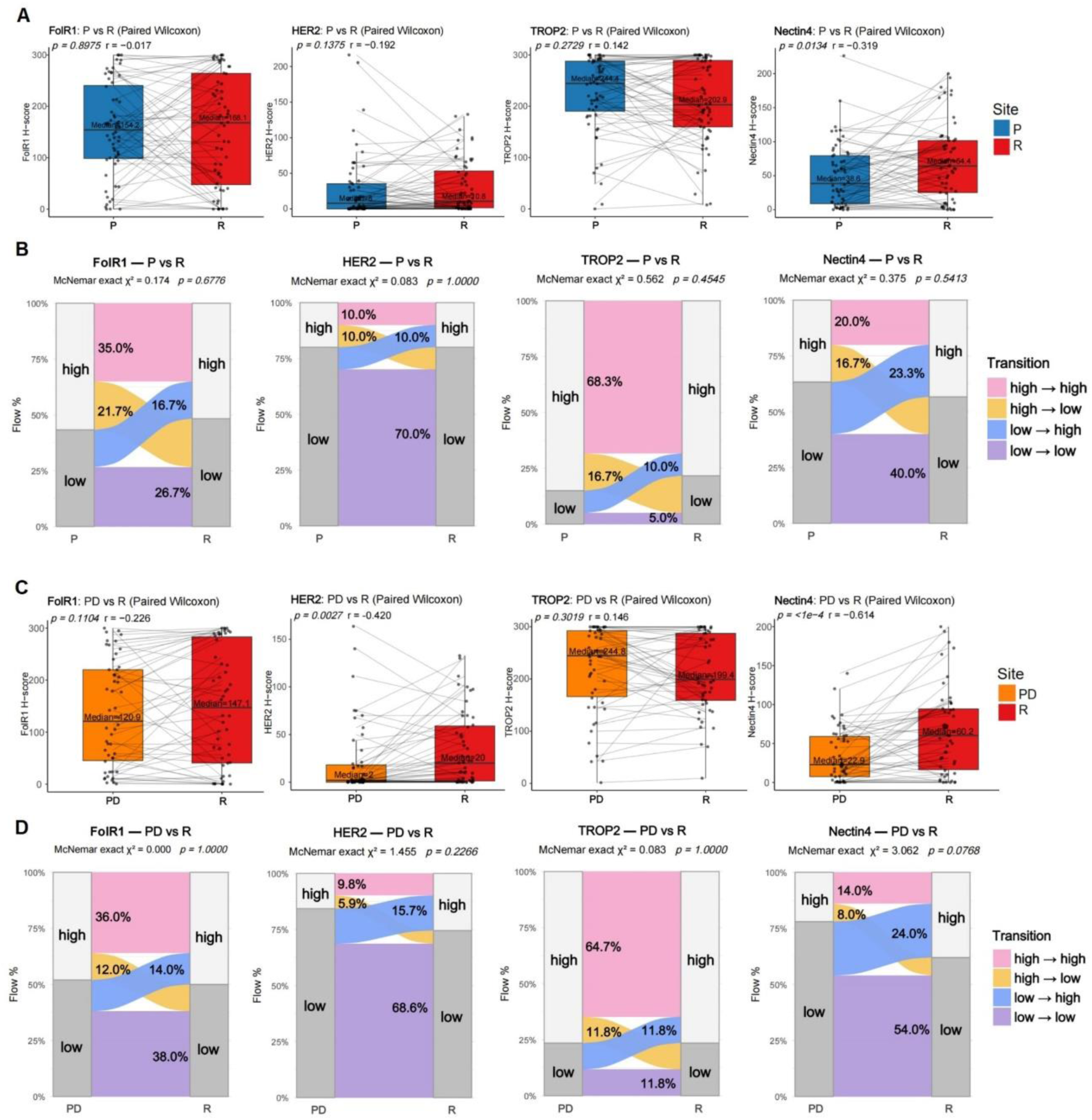
Temporal heterogeneity of ADC target expression in Cohort 2. **(A)** Paired H-score comparisons between P and R samples for FolR1, HER2, TROP2, and Nectin4. Statistical significance was assessed using the paired Wilcoxon test. **(B)** Sankey plots showing categorical group transitions in paired P and R samples across four ADC markers: FolR1, HER2, TROP2, and Nectin4 (left to right). **(C)** Paired H-score comparisons between PD and R samples for FolR1, HER2, TROP2, and Nectin4. Statistical significance was assessed using the paired Wilcoxon test. **(D)** Sankey plots showing categorical group transitions in paired PD and R samples across four ADC markers: FolR1, HER2, TROP2, and Nectin4 (left to right). **Abbreviations**: P, primary ovarian site; PD, primary dissemination; R, recurrence

Second, we compared disseminated primary tumors (PD) with matched recurrent tumors (R) (Fig. 5C–D). H-scores increased in R for Nectin-4 (p<0.0001; r=0.614) and HER2 (p=0.0027; r=0.420), with no significant change for FolR1 or TROP2 (p=0.1104 and p=0.3019, respectively). For binary IHC classifications, category switching was also substantial: Nectin-4 38.0% (McNemar p=0.0768), TROP2 23.6% (p=1.0000), FolR1 26.0% (p=1.0000), and HER2 15.7% (p=0.2268).

These results demonstrate temporal heterogeneity of the ADC target’s expression. At recurrence, H-scores increased for Nectin-4 (vs P and PD) and for HER2 (vs PD). Binary IHC classifications changed in ≥20% of cases across markers, reaching 38–40% for FolR1 and Nectin-4, showing that therapeutic-indication groups can shift over time and should be reassessed at recurrence. The high rate of samples that change expressions between primary and recurrent tumors indicate temporal instability of ADC target expression; reliance on archival primary tissue risks misclassification at recurrence. Clinically, this argues for re-sampling of recurrent, clinically relevant sites with repeat IHC and explicit site/region documentation, as ADC therapy eligibility may change over time.

### 3.4 Prognostic impact of FolR1 expression is site specific

To investigate our hypothesis that ADC marker expression correlates with patient survival, we analyzed the prognostic significance of each marker in Cohort 2, for which we had detailed and long follow-up data. Kaplan–Meier analysis showed that high FolR1 expression in P was significantly associated with worse OS (p = 0.00025) and a borderline association with shorter PFS (p = 0.052) (Fig. 6A). No other ADC markers showed prognostic relevance in P (Supplementary Figure S6). The expression of FolR1 in PD did not show any more association with OS, PFS (Supplementary S7A—B), and in R did not show association with PRS (Fig. 6B). Similarly, the other markers showed a non-significant association with survival in PD and R (Supplementary S7C).

**Figure 6.**
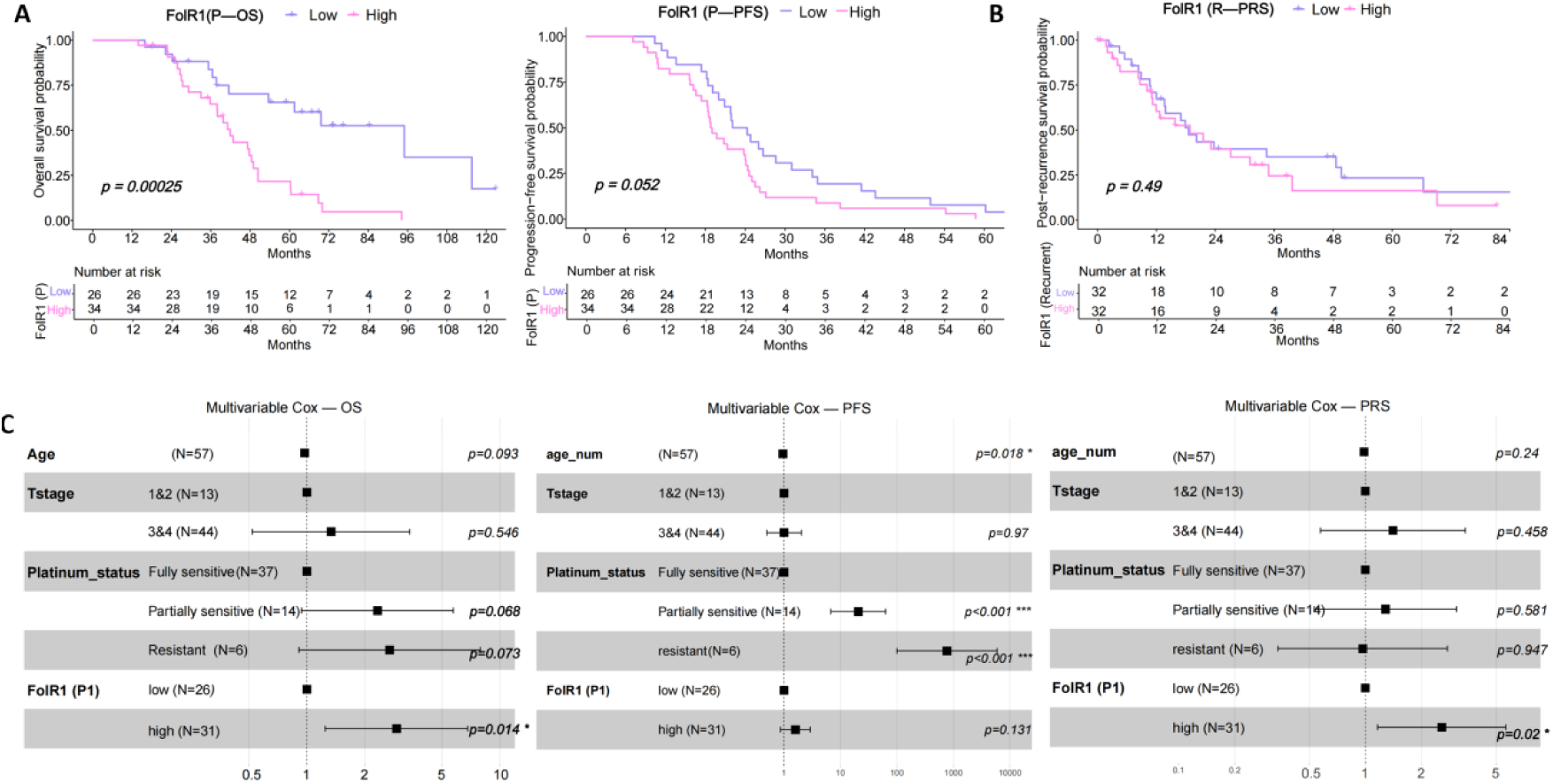
Prognostic impact of FolR1 expression at the primary ovarian site. (**A**) Association of FolR1 expression at P site with OS and PFS in Cohort 2 (Kaplan–Meier). **(B)** Association of FolR1 expression at R site with PRS in Cohort 2 (Kaplan–Meier). **(C)** Independent prognostic value of FolR1 at P for OS, PFS, and PRS (multivariable Cox models adjusted for age, T-stage, and platinum sensitivity). **Abbreviations**: P, primary ovarian site; PD, primary dissemination; R, recurrence; OS, overall survival; PFS, progression-free survival; PRS, post-recurrence survival.

Univariable Cox regression confirmed FolR1 in P as a predictor of poor OS (p = 0.00054). In multivariable Cox regression adjusting for age, T-stage, and platinum sensitivity, FolR1 expression in P remained an independent predictor of OS (p = 0.014), although its association with PFS did not reach significance (p = 0.131) (Fig. 6C). Notably, while FolR1 expression in recurrent tumors was not associated with PRS, multivariable Cox regression indicated that high FolR1 expression at the primary ovarian site was associated with PRS (p=0.02).

These results suggest that FolR1 expression in the primary ovarian site provides robust prognostic information at initial diagnosis, with predictive value that extends to survival even after recurrence, whereas its expression in recurrent tumors itself does not stratify post-recurrence outcomes.

## 4. Discussion

Using two independent TMA cohorts, we comprehensively evaluated molecular, spatial, and temporal heterogeneity of four ADC targets (HER2, TROP2, Nectin-4, FolR1) in HGSOC. We found that: (i) across primary tumors, TROP2 H-scores were lower in *BRCA1/2*-mutated than wild-type tumors (Wilcoxon p=0.0289), while no markers showed significant differences by HRD or *TP53*; most cases showed ≥2 high-expression targets while only weak HER2–Nectin-4 correlation was detected; (ii) spatially, there were no generalizable center–margin or P–PD differences, but the category-switching rates between regions/sites were clinically meaningful, indicating the importance of standardized multi-site and multi-region sampling with IHC; (iii) temporally, Nectin-4 and HER2 tended to be higher at recurrence and FolR1 showed frequent reclassification from P/PD to R, underscoring the need for reassessment; and (iv) high FolR1 expression at the ovarian primary site was associated with worse OS and PRS, whereas its expression in recurrent tumors showed no prognostic value.

FolR1 was frequently high at baseline (∼56%) and showed no molecular heterogeneity. Spatially, FolR1 showed the most marked heterogeneity at P, with 25% center–margin reclassification and also showed notable reclassification between P and PD (21.7%). The binary IHC categories of FolR1 exhibit a high transition rate over time, 38.4% between P and R, and 26% between PD and R—indicating temporal instability. This further underscores the importance of assessing FolR1 expression in the current biopsy, selecting the progressive lesion as the optimal target for sampling. Prior work by Köbel et al. reported an early OS benefit using a broader positivity definition in primary debulking specimens; our stricter cutoff (≥75% moderate/strong) and site-specific analyses (P, PD, R) likely explain the discrepancy. Critically, in our cohort, high FolR1 at P was associated with worse OS and PFS, whereas FolR1 in PD and R was not prognostic. Overall, FolR1 marks baseline aggressiveness and remains a therapeutic relevant target, consistent with the development of MIRV for FolR1-positive platinum-resistant disease^20,23^.

HER2 is largely low in HGSOC, consistent with the historically low rates of HER2 overexpression or amplification^28,47,48^. In our cohorts, HER2 expression is not associated with HRD, *BRCA1/2*, or *TP53* status. Spatially, HER2 showed limited switching: M–C reclassification was 7.2% at P, 11.1% at PD, and 11.2% at R. Between P and PD, only 12.8% of cases changed category. Temporally, HER2 increased at R versus PD (paired Wilcoxon p=0.0027, r=0.42), with binary reclassification in 20.0% cases between P and R and 21.6% between PD and R. Consistently, no association with OS or PFS was observed, in line with Luo et al.’s meta-analysis in serous ovarian cancer^28^.

TROP2 showed the highest prevalence among the ADC targets (∼75–85%). Molecularly, a modest *BRCA1/2*-related decrease in TROP2 H-scores was observed in mutated tumors. Spatially, TROP2 displayed moderate heterogeneity, with 17.9% center–margin switching at P, 24.5% at R, and 23.4% reclassification between P and PD. Temporally, binary categories changed in 26.7% (P–R) and 23.6% (PD–R). Although TROP2 expression was not associated with OS/PFS, its high prevalence across sites and time points indicates reliable target availability, supporting TROP2 as a therapeutic target—in line with the clinical activity of datopotamab deruxtecan (Dato-DXd) in ovarian cancer studies^25–27^.

Nectin-4 showed intermediate prevalence in HGSOC (∼36–37%) and no association with HRD, *BRCA1/2*, or *TP53* status. Spatially, Nectin-4 displayed appreciable heterogeneity, with 19.6% center–margin switching at P and 30.4% reclassification between P and PD—the highest among the four markers. Temporally, Nectin-4 H-scores increased at recurrence and showed frequent binary reclassification (40.0% between P and R; 32% between PD and R). Taken together, Nectin-4 behaves as a dynamic, site- and time-dependent marker in HGSOC. To our knowledge, this is the first systematic evaluation of Nectin-4 in HGSOC, analyzed alongside other ADC targets. Previous studies have not clearly defined its expression or clinical relevance in this tumor type. Given the clinical success of Nectin-4–directed ADC therapy in urothelial carcinoma, although not prognostic in our cohort, its substantial spatial/temporal instability suggests that Nectin-4 may represent a dynamic and therapeutically tractable target in the R setting of HGSOC.TROP2 showed the highest prevalence among the ADC targets (∼75–85%) and no association with HRD, BRCA1/2, or TP53 status. Spatially, TROP2 displayed moderate heterogeneity, with 17.9% center–margin switching at P, 24.5% at R, and 23.4% reclassification between P and PD. Temporally, binary categories changed in 26.7% (between P and R) and 23.6% (between PD and R). Although TROP2 expression was not associated with OS/PFS, its high prevalence across sites and time points indicate reliable target availability, supporting TROP2 as a therapeutic target—in line with the clinical activity of datopotamab deruxtecan (Dato-DXd) in ovarian cancer studies^25–27^.

FDA approval for ovarian cancer therapies often depends on demonstrating a survival benefit. Our data suggest that ADC target expression may have prognostic value, particularly in the primary tumor setting. Similar to *BRCA1/2* as predictive biomarkers for PARP inhibitors, ADC antigen expression might be key to achieving OS benefits. This supports the need for mandatory biomarker-based stratification in ADC trials, ideally through a prospectively defined hierarchical statistical framework.

Our study has several limitations that need to be acknowledged. This study is retrospective and based on TMAs; although each case includes multiple sampling spots, TMAs may still miss subtle spatial heterogeneity observable only in whole-slide images or detectable by spatial omics techniques^49^. We aimed to address this by collecting two regions per tumor — two cores from the center and two from the periphery. This approach has been shown to yield results comparable to whole-slide analysis^50^. Our binary high/low calls relied on predefined cut-offs (e.g., for Nectin-4, thresholds adapted from other tumors without ovarian-specific validation), and category switching rates depend on these cut-offs as well as inter-observer/assay variability. Temporal comparisons may also be influenced by interval length and treatments received between P/PD and R. Sample size limitations reduced the statistical power of certain analyses. Our results support standardized multi-site/multi-region sampling and repeat IHC at recurrence and call for multicenter prospective validation with ovarian-specific cut-offs. Importantly, our study only included patients who were ADC-naïve. As a result, we cannot fully assess how prior exposure to ADCs would alter PFS or OS, nor can we determine how repeated targeting of the same antigen might change the predictive or prognostic value of that antigen over time. This needs to be addressed in future cohorts that include pretreated patients.

ADC targets in HGSOC show limited molecular but marked spatial and temporal heterogeneity. Molecularly, TROP2 H-scores were lower in *BRCA1/2*-mutated cases. No associations were observed among targets or between targets and HRD or *TP53*. The high expression of multiple targets in most cases suggests the feasibility of sequential or alternative targeting strategies. Across targets, we observed clinically meaningful categories switching between regions (M vs C) and sites (P vs PD) and, notably, from P/PD to R. These dynamics indicate that a binary IHC call can change with where and when tissue is sampled. Clinically, this supports standardized multi-site and multi-region sampling, explicit site/region documentation, and repeat IHC at recurrence to minimize misclassification and optimize selection for ADC therapy. In addition, FolR1 at the ovarian primary site marks biologically aggressive disease and retains prognostic value (worse OS and PRS), helping identify patients who may need closer surveillance and informing trial enrollment or treatment choices at diagnosis. Prospective studies should establish ovarian-specific, practical cut-offs and harmonized sampling standards to improve patient selection for ADCs in HGSOC.

## DATA AVAILABILITY

Data underlying the results presented in this paper are not publicly available at this time but may be obtained upon reasonable request to MPD.

## ACKNOWLEDGEMENTS

XL acknowledges support from the China Scholarship Council.

## AUTHOR CONTRIBUTIONS

XL, FD, EIB, and MPD designed the research study. XL and TJ performed experiments. XL, FD, EIB, AD, and MPD collected and annotated clinical data. JS, DH, and WDS supported sample acquisition and logistics. FD, MPD, EIB, MB, SF, and WDS provided critical clinical and pathological input. XL analysed the data. XL, FD, and MPD wrote the manuscript. All authors revised the manuscript and approved the final version.

## FUNDING

This work was supported by the Berlin Institute of Health, Clinician Scientist Program (to MPD), by DKTK Berlin Young Investigator Grant 2022 (to MPD), the German Cancer Aid’s Max-Eder Programme (to FD).

## COMPETING INTERESTS

The authors declare no competing interests.

## ETHICS APPROVAL AND CONSENT TO PARTICIPATE

This study was approved by the local ethics committee (EA1/110/22), with a waiver of patient consent due to the study’s retrospective design and anonymized data. All methods were performed in accordance with the relevant guidelines and regulations.

## CONSENT FOR PUBLICATION

Not applicable. This study uses de-identified retrospective data, and the requirement for patient consent for publication was waived by the ethics boards.

## Notes

### Competing Interest Statement

The authors have declared no competing interest.

### Author Declarations

This study was approved by the Charite Universitaetsmedizin Berlin ethics committee (EA1/110/22), with a waiver of patient consent due to the study's retrospective design and anonymized data.

