## Supplementary table and figures for "Spatial, Temporal, and Molecular Heterogeneity of ADC targets in High-Grade Serous Ovarian Carcinoma"

*Xiaoxuan Li et al.*

Supplementary Table	Page 2

Supplementary Figures	Page 3-9

**Supplementary Table S1.** Clinicopathological characteristics and ADC target expression of two cohorts.

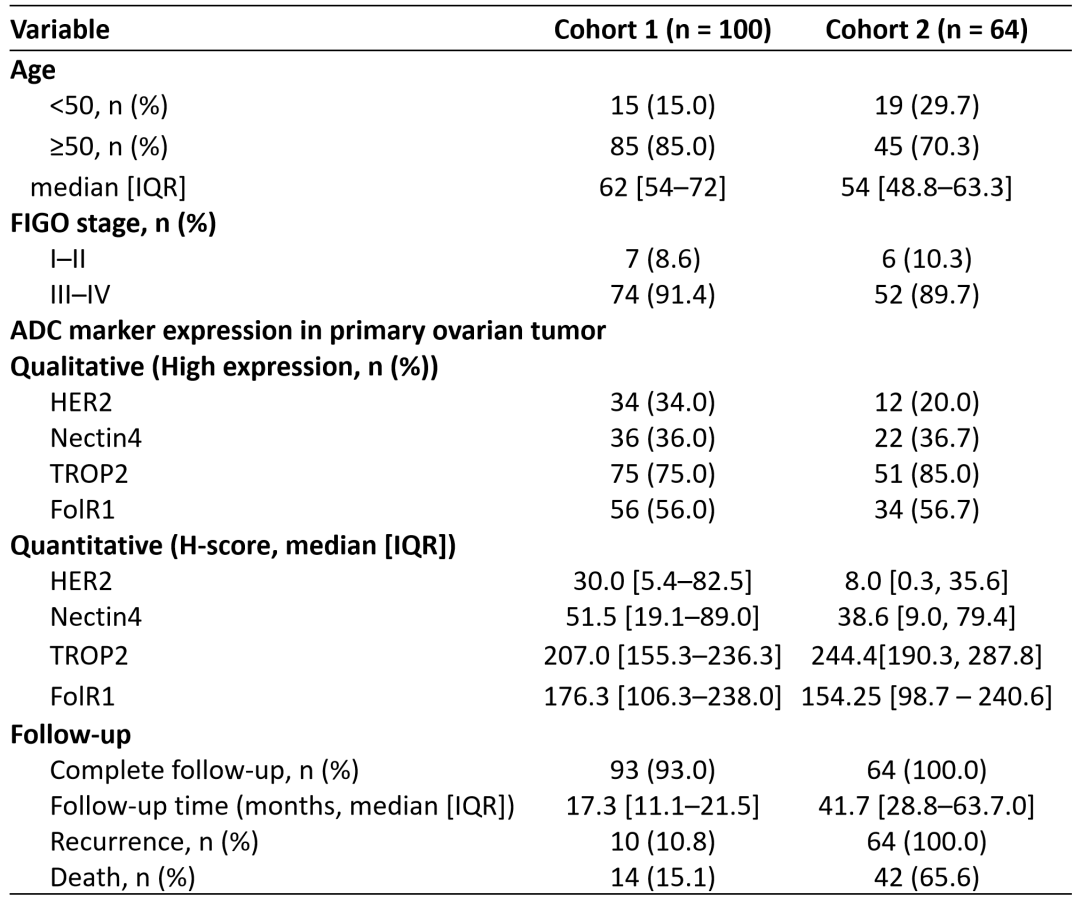

**Abbreviations:** ADC, antibody–drug conjugate; IQR, interquartile range; FIGO, International Federation of Gynecology and Obstetrics; H-score, histological score; HER2, human epidermal growth factor receptor 2; TROP2, trophoblast cell surface antigen 2; FolR1, folate receptor alpha.

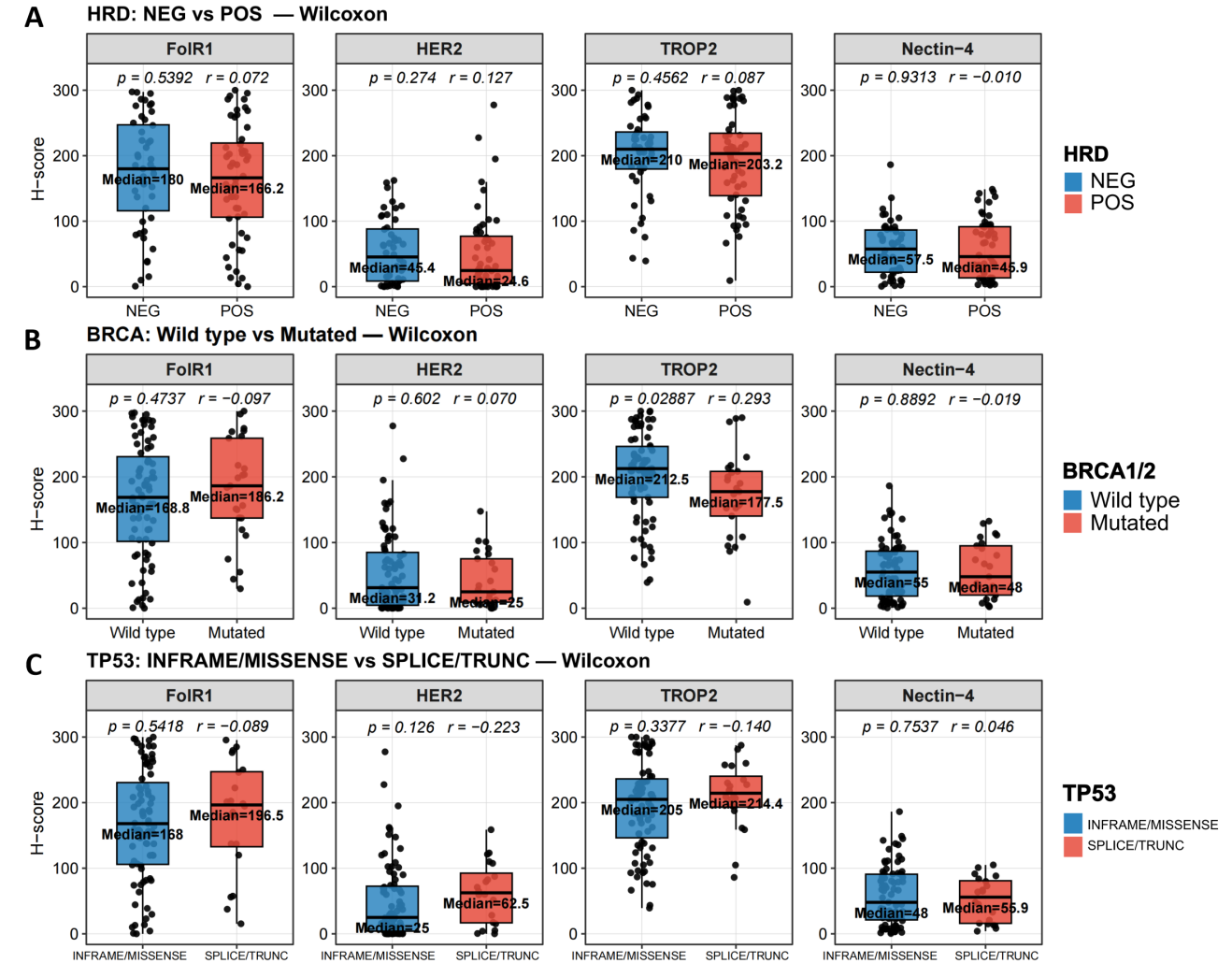

**Supplementary Figure S1. Associations of ADC target expression with genomic features in Cohort 1.**

**(A)** Comparisons between HRD groups (NEG vs POS) for FolR1, HER2, TROP2, and Nectin-4 H-scores.

**(B)** Comparisons between *BRCA1/2* groups (wild type vs mutated) for FolR1, HER2, TROP2, and Nectin-4 H-scores. **(C)** Comparisons between *TP53* mutation classes (in-frame/missense vs splice-site/truncating) for FolR1, HER2, TROP2, and Nectin-4 H-scores. **Abbreviations**: HRD, homologous recombination deficiency; NEG, negative; POS, positive.

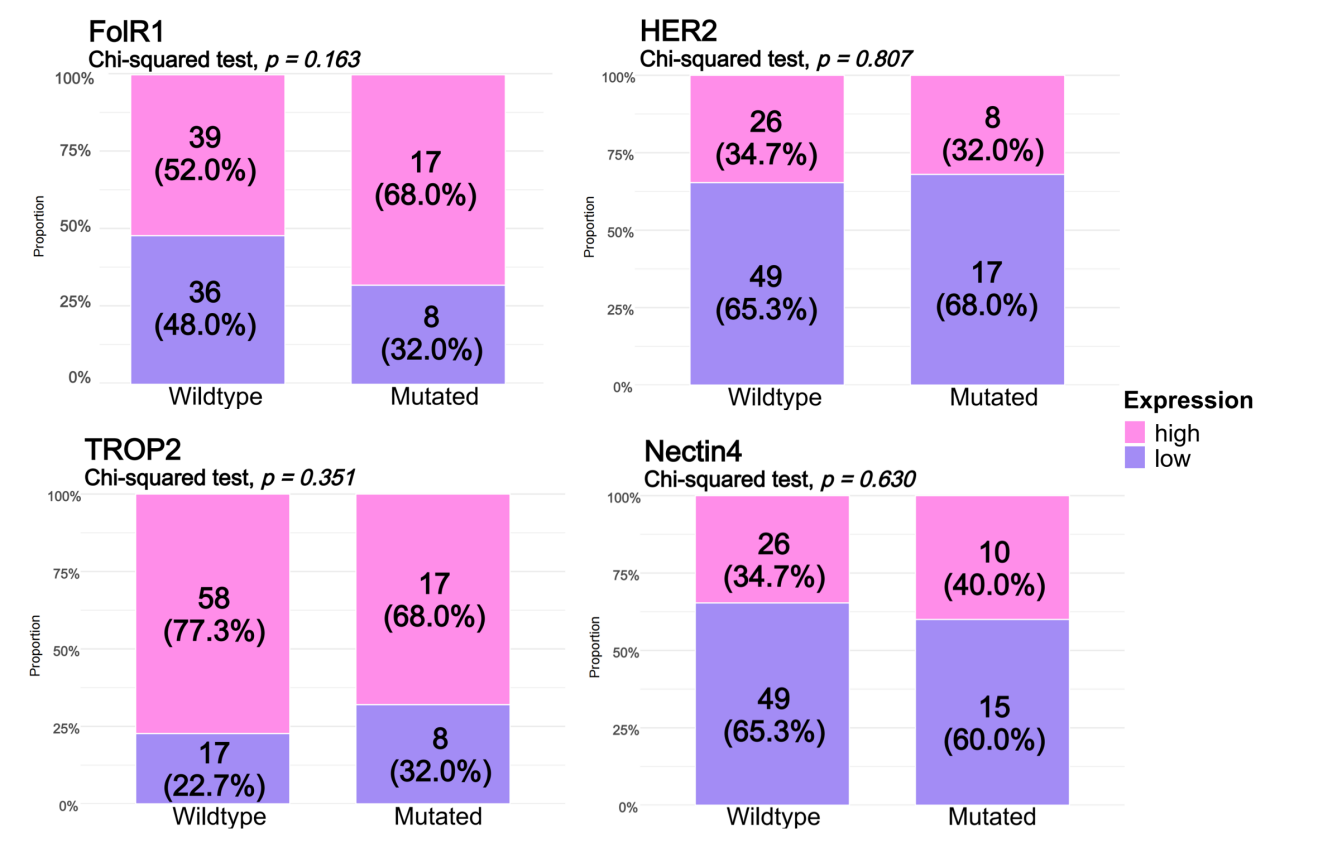

**Supplementary Figure S2. Associations of ADC target expression level with *BRCA1/2* status in Cohort 1.**

Proportion of ADC target expression groups stratified by *BRCA1/2* status (wild type vs mutated) for FolR1, HER2, TROP2, and Nectin-4.

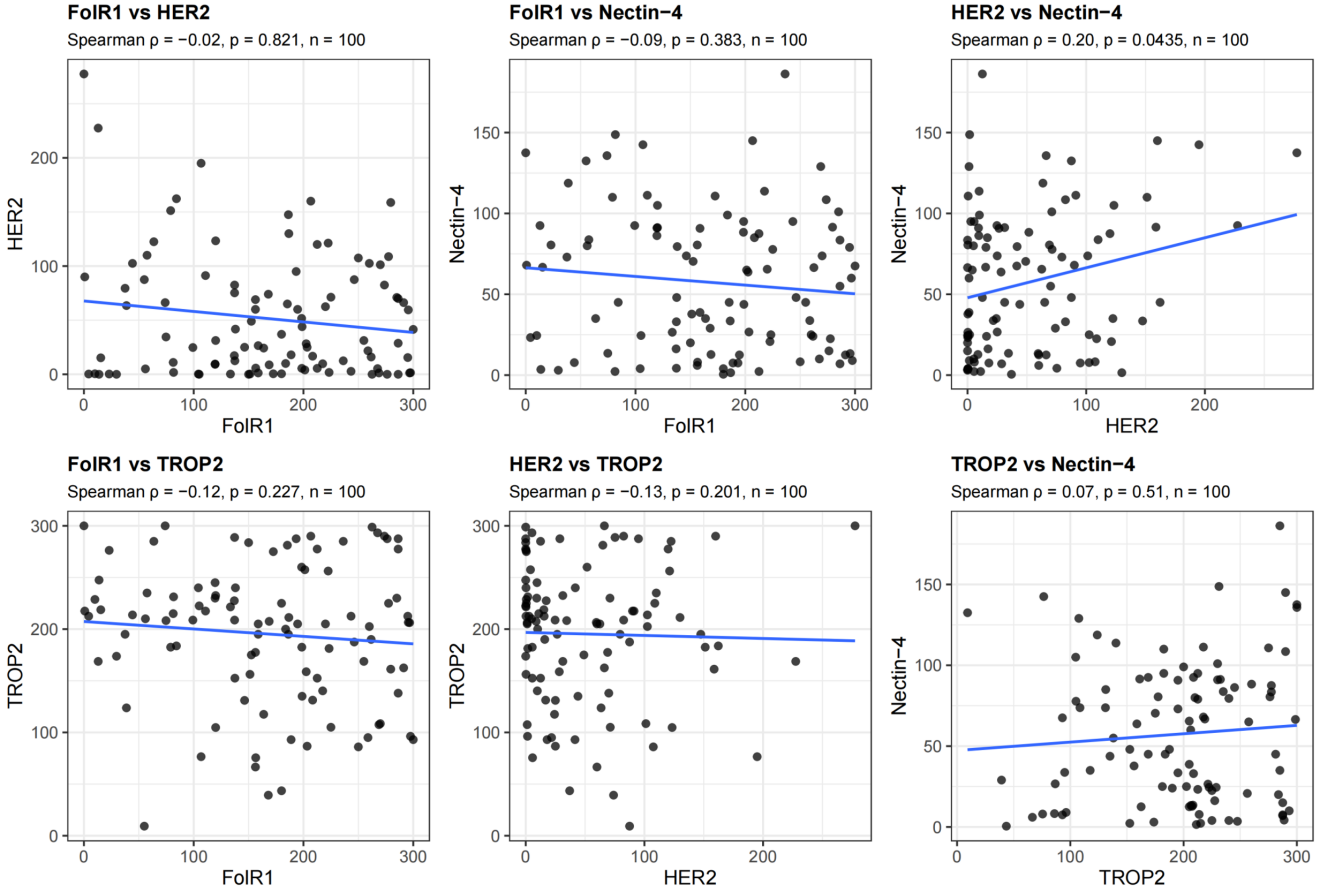

**Supplementary Figure S3. Pairwise correlations among ADC targets in Cohort 1.**

Scatterplots of all pairwise combinations of FolR1, HER2, TROP2, and Nectin-4 H-scores. Each point represents one tumor; the blue line shows a least-squares fit.

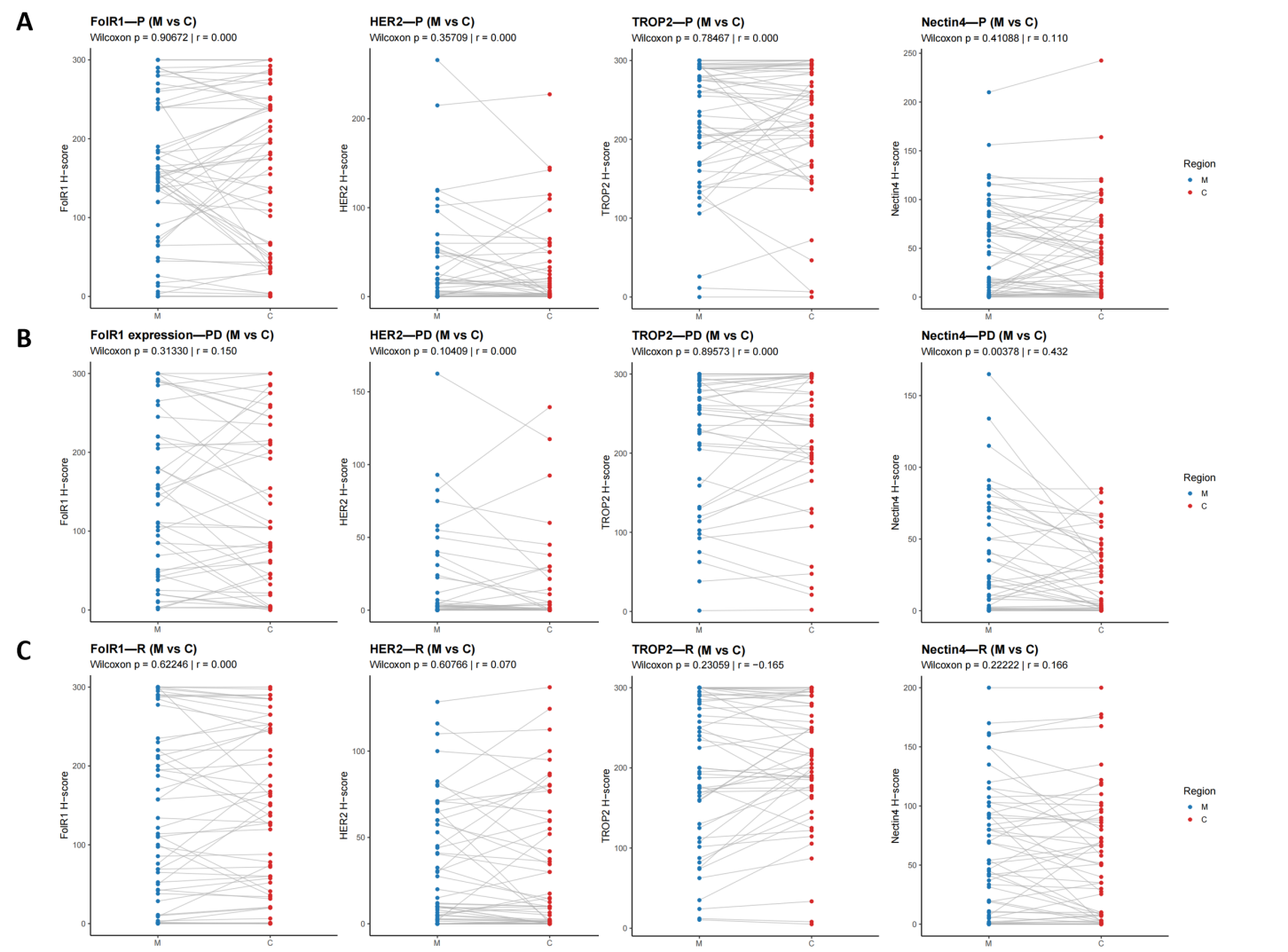

**Supplementary Figure S4. Margin–center comparisons at P, PD, and R across four ADC markers.**

**(A)** Paired H-score comparisons of M versus C for FolR1, HER2, TROP2, and Nectin-4 at P. **(B)** Paired H-score comparisons of M versus C for FolR1, HER2, TROP2, and Nectin-4 at PD. **(C)** Paired H-score comparisons of M versus C for FolR1, HER2, TROP2, and Nectin-4 at R. **Abbreviations**: P, primary ovarian site; PD, primary dissemination; R, recurrent tumor; C, tumor center; M, tumor margin.

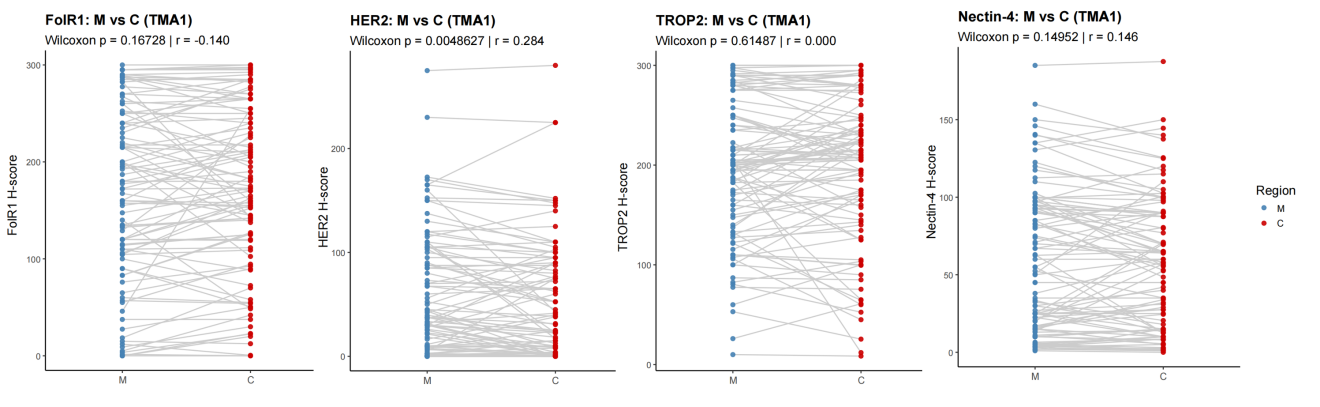

**Supplementary Figure S5. Margin–center comparisons across four ADC markers in Cohort 1.**

Paired H-score comparisons of M versus C for FolR1, HER2, TROP2, and Nectin-4.

**Abbreviations**: C, tumor center; M, tumor margin.

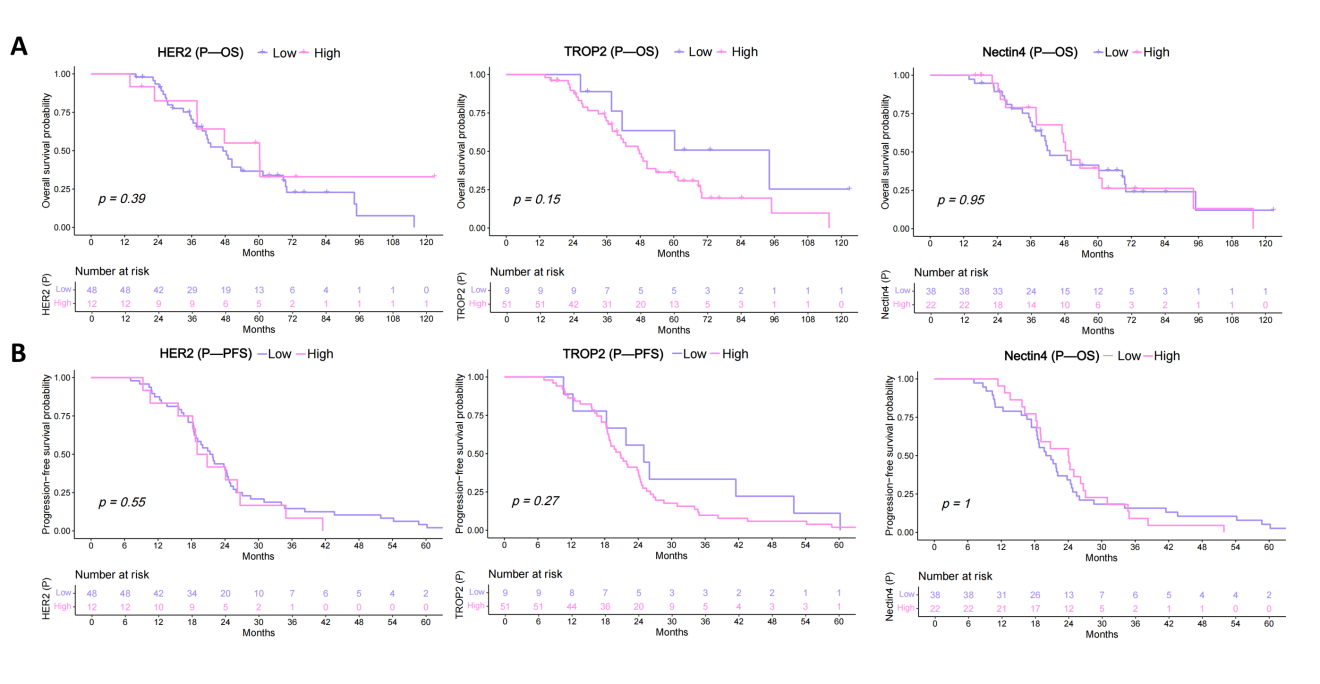

**Supplementary Figure S6. Prognostic analyses at the P site in Cohort 2.**

**(A)** Kaplan–Meier curves comparing OS between high vs low expression groups for HER2, TROP2, and Nectin-4 in P tumors. **(B)** Kaplan–Meier curves comparing PFS between high vs low expression groups for HER2, TROP2, and Nectin-4 in P tumors. **Abbreviations**: P, primary ovarian site; OS, overall survival; PFS, progression-free survival.

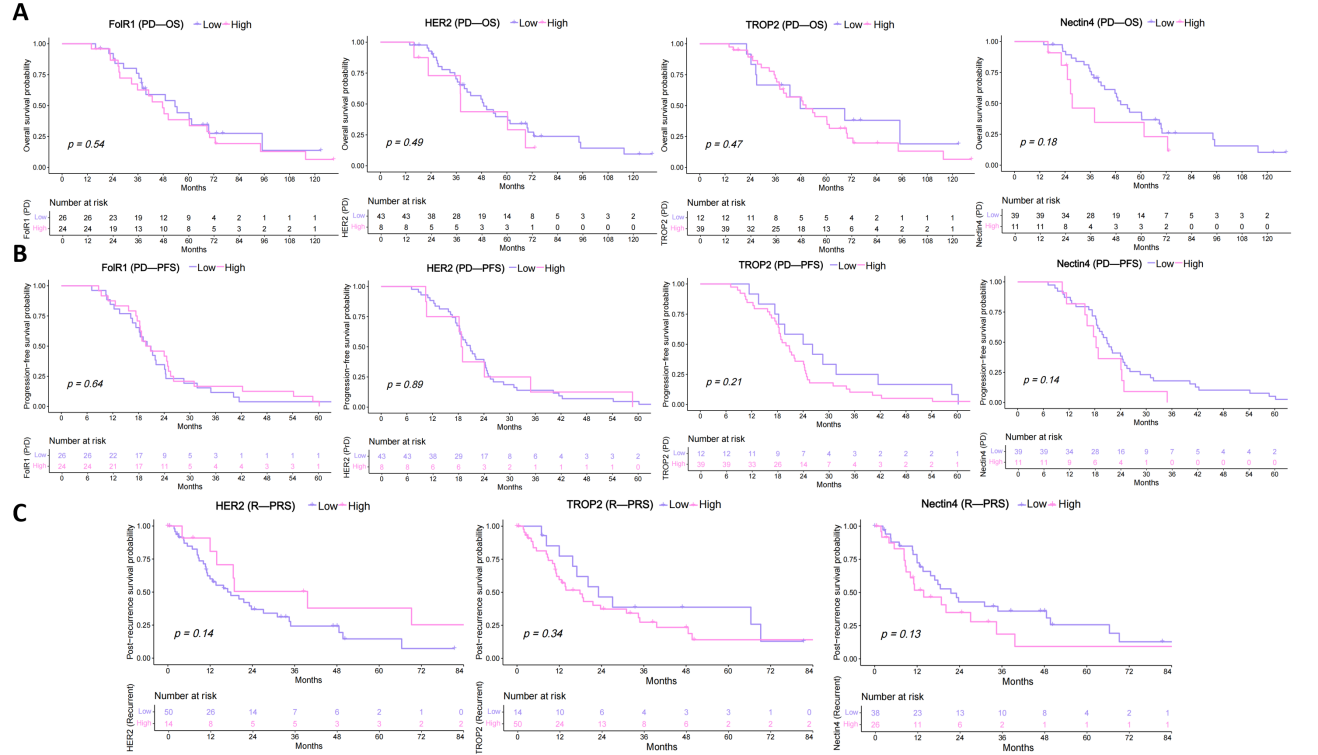

**Supplementary Figure S7. Prognostic analyses at PD and R site in Cohort 2.**

**(A)** Kaplan–Meier curves comparing OS between high vs low expression groups for FolR1, HER2, TROP2, and Nectin-4 in PD tumors. **(B)** Kaplan–Meier curves comparing PFS between high vs low expression groups for FolR1, HER2, TROP2, and Nectin-4 in PD tumors. **(C)** Kaplan–Meier curves comparing PRS between high vs low expression groups for HER2, TROP2, and Nectin-4 in PD tumors. **Abbreviations**: PD, primary dissemination; R, recurrent tumor; OS, overall survival; PFS, progression-free survival; PRS, post-recurrence survival.
